# A Proof-of-Concept Study of a Clinical Decision Support System for Vancomycin Therapeutic Monitoring

**DOI:** 10.64898/2026.02.22.26346368

**Authors:** Fahmi Hassan, Lou Jing Ying, Lim Chong Teck, Ong Wei Quan, Nurul Nasuhah Rumaizi

## Abstract

Artificial intelligence (AI), particularly large language models (LLMs), is increasingly explored in healthcare, yet its real-world usability and safety in high-risk clinical pharmacy tasks remain uncertain. Vancomycin therapeutic drug monitoring (TDM), which requires precise pharmacokinetic calculations and context-sensitive interpretation within a narrow therapeutic window, provides a stringent test case for AI-assisted decision support. This proof-of-concept study developed and evaluated a hybrid clinical decision support system (TDM-AID) integrating a validated deterministic pharmacokinetic calculation engine, GPT-4o–based structured clinical interpretation, and retrieval-augmented guideline support. Thirty retrospective adult vancomycin TDM cases were assessed using a weighted six-domain rubric covering pharmacokinetic accuracy, AUC estimation, prospective prediction, timing recommendations, clinical judgment, and documentation quality. Two independent expert pharmacists evaluated system outputs against benchmark consultations. The overall median performance was 78% (IQR 12%), classified as Acceptable, and 73% (IQR 14%) when deterministic calculations were excluded. Foundational pharmacokinetic calculations achieved 100% accuracy. Clinical judgment demonstrated Good performance (83%), whereas prospective prediction was limited (58%), and timing recommendations were absent in all cases. Safety violations occurred in 17% of cases, including dose recommendations exceeding 4 g/day. Inter-rater reliability was good (ICC 0.87). These findings suggest that hybrid AI-driven decision support is technically feasible and usable as a pharmacist-augmenting draft generator; however, limitations in predictive reasoning, timing logistics, and safety enforcement necessitate deterministic safeguards and mandatory expert oversight before clinical implementation.

## 1. Introduction

Therapeutic drug monitoring (TDM) involves individualization of drug dosages to maintain blood concentrations within targeted therapeutic ranges, ensuring both efficacy and safety[1]. This approach is particularly critical for drugs with narrow therapeutic windows in which small deviations can result in treatment failure or serious adverse effects[2]. Vancomycin, a glycopeptide antibiotic used for serious gram-positive infections including methicillin-resistant Staphylococcus aureus (MRSA), exemplifies medications requiring careful monitoring due to dose-dependent nephrotoxicity[3, 4]. Updated consensus guidelines from the American Society of Health-System Pharmacists (ASHP), Infectious Diseases Society of America (IDSA), and Society of Infectious Diseases Pharmacists (SIDP) now recommend area under the concentration-time curve over 24 hours (AUC_24_) as the primary pharmacokinetic parameter for vancomycin TDM, with target ranges of 400-600 mg·h/L for most serious MRSA infections[5]. This shift from trough-based to AUC-based monitoring requires Bayesian estimation or two-point pharmacokinetic modeling, which adds to pharmacist workload[6, 7].

Malaysian hospital pharmacies face substantial challenges delivering comprehensive TDM services, with high patient volumes and staffing constraints limiting time available for detailed pharmacokinetic calculations, clinical assessments, and dosing recommendations[8]. The complexity of AUC-based monitoring further increases cognitive burden on clinical pharmacists[9]. These operational constraints may lead to delays in TDM consultations, potentially compromising patient outcomes and creating demand for workflow optimisation solutions[8, 10]. Recent advances in artificial intelligence have enabled the development of clinical decision support systems (CDSS) combining computational power with clinical guideline adherence[11, 12]. Large language models (LLMs) such as GPT-4o show potential for processing complex clinical data and generating structured recommendations. Studies examining ChatGPT in clinical pharmacy tasks found 100% success in identifying drug interactions, though with limitations in specific dosage recommendations[13]. Machine learning approaches to vancomycin dosing have demonstrated promising target attainment rates comparable to expert recommendations[14]. However, most existing AI applications focus narrowly on dose prediction and do not address the broader consultation workflow, including AUC calculation, contextual clinical interpretation, and structured documentation[15]. Furthermore, unconfigured large language models have demonstrated inconsistent reliability in therapeutic drug monitoring tasks, underscoring the need for controlled implementation strategies[16].

Collectively, these findings suggest that the challenge is not solely dose prediction, but integration. Deterministic pharmacokinetic computation provides mathematical precision, whereas large language models offer capacity for structured synthesis of clinical data. When coupled with retrieval-augmented generation to anchor outputs in established guidelines, a hybrid architecture may address both computational accuracy and interpretive consistency. This study aimed to develop and validate a hybrid clinical decision support system for vancomycin therapeutic drug monitoring that integrates deterministic pharmacokinetic modeling with large language model–assisted clinical interpretation. System performance was evaluated in terms of computational accuracy and concordance of clinical recommendations against reference standards.

## 2. Methods

### 2.1 Study Design and Setting

A retrospective, cross-sectional proof-of-concept study was conducted at a tertiary care hospital in Malaysia. The study consisted of two sequential phases: (1) system development and technical configuration, followed by (2) performance evaluation using real-world vancomycin therapeutic drug monitoring (TDM) cases.

### 2.2 System Development (TDM-AID)

TDM-AID was designed as a three-module hybrid architecture executed sequentially. The first module comprised deterministic pharmacokinetic calculation functions implementing first-order elimination equations for vancomycin. Calculated parameters included elimination rate constant (ke), half-life (t½), volume of distribution (Vd), clearance (CL), and area under the concentration–time curve over 24 hours (AUC_24_). All equations were implemented using established pharmacokinetic formulas[17, 18]. Outputs were verified against manual reference calculations to confirm computational accuracy and numerical stability.

The second module employed GPT-4o (OpenAI) as the clinical interpretation engine to process computed pharmacokinetic parameters and generate structured dosing recommendations. The model was accessed via API integration and configured using structured prompts to standardize output format. Model selection was based on its demonstrated performance in medical knowledge tasks[19], ability to process extended clinical context within a single interaction[20], and compatibility with structured API-based deployment required for system integration[19]. The third module implemented retrieval-augmented generation (RAG) using the Clinical Pharmacokinetics Pharmacy Handbook[21]. Relevant guideline sections were dynamically retrieved and supplied to the language model to constrain responses to locally endorsed practice standards and reduce unsupported output generation.

The clinical interpretation module was configured through systematic prompt engineering. A structured input template provided patient demographics, renal function parameters (serum creatinine, creatinine clearance), current vancomycin regimen details, measured drug concentrations with precise timing, infection characteristics, and target therapeutic parameters. The system-level prompt established the role as a clinical pharmacist specialist in antimicrobial stewardship and pharmacokinetics. Task-specific prompts provided patient information, pre-calculated pharmacokinetic parameters, and relevant clinical guidelines extracted through RAG. The prompt instructed the interpretation module to assess current dosing and drug levels, evaluate appropriateness of calculated doses, identify factors necessitating adjustments beyond basic calculations, provide specific guideline-tied rationale, and generate monitoring recommendations formatted as concise clinical consultation notes.

Model parameters included temperature 0.2 to promote consistency, a maximum of 1500 tokens, and default top-p sampling. Low temperature prioritized deterministic, guideline-adherent recommendations over creative interpretation. The RAG system processed guideline PDFs into text chunks (1000 characters with 200-character overlap), embedded them using text-embedding-3-small, and indexed using the FAISS vector database. For each case, the top three most relevant guideline excerpts were retrieved based on semantic similarity to queries incorporating patient-specific factors. The interpretation module received pre-calculated parameters from the validated engine and was instructed to interpret pharmacokinetic results relative to therapeutic targets, compare against guideline-derived ranges, generate dosing recommendations achieving target AUC_24_ (400–600 mg·h/L for empirical therapy, higher for definitive therapy based on organism characteristics)[22], predict prospective concentrations following recommended changes, provide resampling timing and monitoring guidance, and document findings with guideline citations. The complete application source code is provided in Appendix A for transparency and reproducibility.

### 2.3 Performance Assessment

Thirty retrospective vancomycin TDM cases were selected via purposive sampling from pharmacy consultation records (January 2022 to December 2024). Inclusion criteria required adult patients (≥18 years) with complete clinical data including demographics, renal function, dosing history, and measured vancomycin concentrations. Cases included patients with normal or stable renal function where standard pharmacokinetic calculations are reliable. Exclusion criteria encompassed highly unstable renal function (acute kidney injury with rapidly changing creatinine, renal replacement therapy), initial dose consultations without subsequent measured levels, and incomplete documentation.

The final sample represented both definitive therapy (target AUC_24_ >600 mg·h/L) and empirical therapy (target AUC_24_ 400-600 mg·h/L) scenarios across various infection sites. Follow-up TDM cases for the same patients were included as they represent realistic clinical scenarios requiring consideration of trends and evolving patient status. A comprehensive assessment rubric was developed through systematic review of TDM practice standards and clinical pharmacy competency frameworks[23]. Six domains were identified as essential for vancomycin TDM consultation. Domain A (foundational calculations, weight 2×) assessed basic pharmacokinetic parameters. Domain B (parameter estimation and AUC, weight 3×) evaluated current exposure assessment. Domain C (prospective predictions, weight 3×) examined prediction of future steady-state concentrations. Domain D (timing recommendations, weight 2×) focused on resampling guidance. Domain E (interpretation and core recommendations, weight 5×) assessed clinical judgment, dose appropriateness, and safety considerations. Domain F (clinical reporting, weight 1×) evaluated report clarity and consistency. The complete evaluation rubric used for scoring is provided in Appendix B.

Weights reflected relative importance to patient safety and clinical decision-making based on expert consensus. Domains with direct impact on dosing decisions received higher weights, while foundational calculations received moderate weighting as they serve as prerequisites but rarely vary when performed correctly. Each rubric item was scored on a three-point scale: 0 for inadequate or incorrect, 1 for partially correct, and 2 for fully correct or optimal. An overarching safety violation category captured recommendations posing significant patient harm, triggering substantial score reductions. Two independent expert pharmacists with extensive TDM experience (>5 years) evaluated each TDM-AID output using the assessment rubric after standardized training. Both reviewers compared system outputs against original pharmacist-completed consultations. Rubric items were scored for each domain, with scores excluded when not applicable. Data normalization was performed by calculating the percentage of the maximum possible score for each domain.

Inter-rater reliability was assessed using intraclass correlation coefficient (ICC) with a two-way random effects model for absolute agreement[24, 25]. ICC interpretation followed established guidelines: less than 0.50 indicates poor reliability, 0.50-0.75 moderate, 0.75-0.90 good, and greater than 0.90 excellent reliability[25]. Mean scores from both reviewers were used for final analyses. Statistical analyses were performed using Microsoft Excel and Python (SciPy statistical package). Descriptive statistics were reported as median and interquartile range (IQR) for domain-specific and total percentage scores. Overall system performance was calculated across all six domains using weighted scoring based on predefined clinical importance. A sensitivity analysis was conducted excluding Domain A (deterministic pharmacokinetic calculations) to evaluate performance in clinically interpretive domains, thereby isolating the contribution of the AI-assisted components. Performance was categorized using predefined thresholds as follows: Excellent (90–100%), Good (80–89%), Acceptable (70–79%), and Needs Improvement (<70%). Data distribution was assessed using the Shapiro–Wilk test. As normality assumptions were not consistently met, paired comparisons between TDM-AID outputs and pharmacist benchmark assessments were performed using the Wilcoxon signed-rank test. All statistical tests were two-tailed, and statistical significance was defined as p < 0.05.

### 2.4 Ethical Considerations

This study was approved by the Medical Research and Ethics Committee (MREC), Ministry of Health Malaysia. The study utilized retrospective, de-identified patient data for evaluation of a clinical decision support tool. As the research involved analysis of existing clinical records without direct patient contact or intervention, informed consent was waived in accordance with MREC approval and national ethical guidelines.

## 3. Results

The overall median performance score of TDM-AID was 78% (IQR 12%), classified as Acceptable. Sensitivity analysis excluding Domain A (foundational pharmacokinetic calculations) yielded a median score of 73% (IQR 14%), also classified as Acceptable. In the primary analysis, 53% of cases (n=16) achieved Acceptable performance (70–79%), with the remaining cases distributed across Good (80– 89%), Excellent (90–100%), and Needs Improvement (<70%) categories. Detailed domain-specific scores and statistical comparisons are summarized in Table 1. The validated pharmacokinetic calculation engine (Domain A) achieved 100% performance across all 30 cases (median 100%, IQR 0%) for elimination rate constant, volume of distribution, clearance, and half-life calculations. No discrepancies were identified. Statistical comparison with pharmacist benchmarks was not performed due to identical performance. For Parameter Estimation and AUC (Domain B), the median score was 100% (IQR 0%). There was no statistically significant difference compared with pharmacist benchmarks (Wilcoxon signed-rank p=0.066).

**Table 1.** Performance Scores of TDM-AID Across Individual and Aggregate Domains Compared to Pharmacist Benchmarks.

| Domain | Median (IQR) | Classification |
| --- | --- | --- |
| Overall (All Domains) | 78% (12%) | Acceptable |
| Clinical Domains (B-F) | 73% (14%) | Acceptable |
| A. Foundational Calculations | 100% (0%) | Excellent |
| B. Parameter Estimation & AUC | 100% (0%) | Excellent |
| C. Prospective Predictions | 58% (17%) | Needs improvement |
| D. Timing Recommendations | NA | NA |
| E. Clinical Judgement | 83% (25%) | Good |
| F. Documentation Quality | 71% (15%) | Acceptable |

For Prospective Predictions (Domain C), the median score was 58% (IQR 17%), classified as Needs Improvement. Performance differed significantly from pharmacist benchmarks (Wilcoxon signed-rank p<0.05). Discrepancies included inaccurate steady-state projections and inconsistencies between recommended dose adjustments and predicted pharmacokinetic outcomes. The system did not provide specific resampling dates or times under Timing Recommendations (Domain D) in any of the 30 cases. As no cases met scoring criteria for this domain, descriptive and inferential statistics were not calculated. For Clinical Judgment and Core Recommendations (Domain E), the median score was 83% (IQR 25%), classified as Good. Performance differed significantly from pharmacist benchmarks (Wilcoxon signed-rank p<0.05). Documentation Quality (Domain F) demonstrated a median score of 71% (IQR 15%), classified as Acceptable. Performance differed significantly from pharmacist benchmarks (Wilcoxon signed-rank p<0.05). Observed issues included inconsistencies in dosing statements and variable clarity of recommendations. Safety-related concerns were identified in five cases (17%) where recommended vancomycin doses exceeded 4 g/day. These cases were classified under the Overarching Safety Violations category and received substantial score reductions. Three additional cases (10%) involved dosing frequency recommendations inconsistent with calculated vancomycin half-life.

Inter-rater reliability demonstrated good to excellent agreement between independent reviewers. The overall intraclass correlation coefficient (ICC) for total percentage scores was 0.87 (95% CI: 0.71– 0.95). Domain-specific ICC values ranged from 0.92 to 1.00 for calculation-based domains. For domains requiring clinical judgment, ICC values were 0.81 (95% CI: 0.62–0.92) for Domain C, 0.79 (95% CI: 0.58–0.91) for Domain E, and 0.74 (95% CI: 0.49–0.88) for Domain F.

## 4. Discussion

This study evaluated the feasibility of TDM-AID, a hybrid clinical decision support system integrating validated pharmacokinetic calculations, LLM-based clinical interpretation, and retrieval-augmented guideline support for vancomycin TDM. The system achieved perfect accuracy in deterministic pharmacokinetic calculations while demonstrating variable performance in clinical reasoning domains. Overall performance of 78% (73% when excluding deterministic calculations) supports feasibility, but identifies optimization priorities in prospective prediction (58%), timing recommendations (absent), and documentation consistency (71%).

While the overall median performance of 78% is classified as “Acceptable” under the predefined grading framework, this classification must be interpreted with caution in a clinical context. Vancomycin has a narrow therapeutic window, where relatively small dosing deviations may result in subtherapeutic exposure or dose-dependent nephrotoxicity[3, 32]. In this study, 17% of cases involved safety violations, specifically recommendations exceeding 4 g/day. Such a rate is clinically unacceptable for any autonomous or unsupervised system. Although the weighted rubric categorizes overall performance as acceptable, patient safety considerations supersede numerical grading. These findings reinforce that TDM-AID is intended strictly as a draft-generating and decision-support tool to augment expert pharmacist review. It is not designed for independent clinical deployment. The strength of the deterministic calculation module lies in reducing manual computational workload; however, interpretive outputs generated by a probabilistic language model require mandatory verification by a qualified clinician. This distinction reflects broader implementation science principles emphasizing human oversight when integrating generative AI into high-risk healthcare workflows[31].

The validated calculation engine achieved 100% accuracy across all foundational pharmacokinetic parameters. This confirms the architectural advantage of separating deterministic computation from probabilistic language-model interpretation. Arithmetic operations, unit conversions, and first-order kinetic equations are inherently rule-based and benefit from validated mathematical functions rather than LLM generation[26]. This design leverages the distinct characteristics of rule-based and probabilistic reasoning described in clinical decision support literature[27]. The computational layer therefore represents a reliable and immediately deployable component of the system. In contrast, performance declined in domains requiring clinical reasoning. Large language models operate through probabilistic modeling rather than causal inference[28]. Clinical pharmacokinetics requires anticipatory reasoning—predicting how dose modifications alter steady-state exposure under patient-specific conditions. The reduced performance in prospective prediction suggests that free-text LLM extrapolation is insufficient for forecasting concentration changes after dose adjustment. Similar limitations have been reported in evaluations of LLM performance managing complex medication regimens[16]. These findings support integration of specialized prediction algorithms or Bayesian forecasting tools within the hybrid framework rather than reliance on generative reasoning alone.

A notable limitation was the complete absence of output in Domain D (Timing Recommendations), resulting in 0% performance for resampling guidance. In all 30 cases, the system failed to provide specific resampling dates or times. This reflects a compounded limitation of prompt design and model constraint rather than isolated omission. Although system-level instructions requested monitoring guidance and resampling timing, the model did not translate this directive into explicit, operational schedules. This finding highlights a structural limitation of relying on general LLM reasoning for time-sensitive clinical logistics. Determining optimal sampling windows is algorithmic and dependent on elimination half-life, dosing interval, and attainment of steady state—variables that are mathematically definable rather than interpretive. Future iterations should therefore bypass the LLM for this function and implement a deterministic timing algorithm that calculates resampling windows directly from pharmacokinetic parameters and dosing schedules. Such modular replacement aligns with hybrid CDSS design principles that allocate structured tasks to rule-based engines while reserving narrative synthesis for LLM components[27].

Discrepancies in clinical judgment further reflect differences between statistical language modeling and experiential pharmacist reasoning. Expert clinicians integrate contextual factors such as infection severity, nephrotoxicity risk, microbiological data, and institutional practice patterns. Such multidimensional reasoning is difficult to encode fully within structured prompts. Instances where the system recommended dose escalation despite attainment of target AUC_24_ illustrate sensitivity to how therapeutic thresholds are framed. This observation aligns with evidence that LLM outputs vary significantly based on prompt structure and framing[29]. Safety findings further underscore the need for architectural guardrails. Recommendations exceeding safe dosing limits are consistent with documented vulnerabilities of generative models, including production of plausible yet clinically inappropriate recommendations when extrapolating beyond reliable distributions[30]. In safety-critical domains such as antimicrobial TDM, hard-coded dose ceilings, interval validation against calculated half-life, and automated conflict checks must override generative outputs when predefined safety thresholds are exceeded. Governance-oriented implementation frameworks emphasize this requirement for structural constraints in healthcare AI deployment[31].

Interpretation of therapeutic targets also contributed to discrepancies. Contemporary consensus guidelines recommend AUC_24_ targets of 400–600 mg·h/L for most serious MRSA infections to balance efficacy and nephrotoxicity risk[5, 32]. Evidence linking higher exposures to increased acute kidney injury further supports strict adherence to this range[9]. These findings illustrate that AI-assisted systems are highly sensitive to explicit framing of therapeutic targets and curated reference materials supplied through retrieval augmentation. Comparison with prior literature situates these findings within the broader AI-in-pharmacy context. General-purpose LLMs have demonstrated capability in identifying drug interactions[13, 33], while machine learning models tailored for vancomycin dosing prediction show improved initial dose estimation accuracy[14, 34]. However, most prior work evaluates isolated predictive tasks rather than full consultation workflows. The present system attempted comprehensive TDM consultation—calculation, interpretation, prediction, monitoring, and documentation—thereby exposing variability across domains. The structured, weighted rubric applied in this study contributes methodological transparency to AI performance evaluation in pharmacy practice[35].

The findings define clear development priorities. Prospective prediction should transition from LLM-based extrapolation to validated forecasting engines or Bayesian tools. Deterministic timing algorithms must replace generative approaches for resampling guidance. Architectural safety constraints should enforce maximum dose limits and pharmacokinetic consistency checks. Structured documentation templates and validation checks may further improve reporting clarity. These results support positioning TDM-AID as a pharmacist-augmenting system rather than an autonomous decision-maker. Immediate applications include automated calculation support and generation of draft consultation notes for pharmacist review. Current safety findings do not support unsupervised deployment. A phased development approach is warranted, beginning with architectural refinements and followed by supervised prospective evaluation. Expansion to populations with unstable renal function, renal replacement therapy, pediatric pharmacokinetics, or critical illness will require further validation. Parameter-efficient fine-tuning using institutional TDM datasets may improve contextual alignment while maintaining governance standards. Federated learning models may facilitate multi-center improvement without compromising data privacy[36].

Several limitations require acknowledgment. The study included only adult patients with stable renal function, limiting generalizability. Pharmacist consultations were used as reference standards, assuming but not guaranteeing optimality. Moderate inter-rater variability in subjective domains reflects inherent differences in clinical reasoning. The rubric, while systematically developed, was not externally validated. Only one LLM architecture was assessed, and performance may differ across models. Overall, this proof-of-concept demonstrates that hybrid architectures combining deterministic pharmacokinetic engines with LLM-assisted interpretation are technically feasible and computationally reliable. However, acceptable numerical performance does not equate to autonomous clinical safety. Robust guardrails, deterministic algorithms for structured tasks, and mandatory expert oversight remain essential prerequisites for responsible clinical integration.

## 5. Conclusion

This study demonstrates that TDM-AID, a hybrid system combining validated pharmacokinetic calculators with LLM-based interpretation and RAG-enabled guideline retrieval, achieves perfect computational accuracy while highlighting areas for refinement in clinical interpretation (73% performance excluding calculations). Key limitations include prospective predictions, timing recommendations, and documentation consistency, which can be addressed through safety constraints, specialized prediction algorithms, deterministic timing logic, and structured documentation templates. A phased development approach with mandatory pharmacist review and prospective evaluation is recommended to ensure patient safety. TDM-AID provides a feasible framework for augmenting hospital pharmacy TDM workflows, enhancing pharmacist decision-making while preserving essential clinical judgment.

## Supporting information

Apendix A

## Acknowledgments

The authors thank the pharmacy department of Hospital Tengku Ampuan Rahimah and therapeutic drug monitoring team for their support in conducting this research. We acknowledge the contributions of the TDM-specialized pharmacists whose expert consultations served as the benchmark for this evaluation.

## Conflict of Interest

The authors declare no conflicts of interest related to this research.

## Funding

This research received no specific grant from any funding agency in the public, commercial, or not-for-profit sectors.

## Data Availability

De-identified data supporting the findings of this study are available from the corresponding author upon reasonable request and subject to institutional data sharing policies.

## Author Contributions

Conceptualization: FH Methodology: FH, LJY, LCT

Software (System Development): FH Formal Analysis: OWQ, NNR, FH

Investigation (Data Acquisition): OWQ, NNR, LJY, LCT

Data Curation: OWQ, NNR, LJY, LCT

Writing – Original Draft Preparation: OWQ, NNR

Writing – Review & Editing: FH, LJY, LCT

Supervision: FH

Project Administration: FH

## Notes

### Competing Interest Statement

The authors have declared no competing interest.

### Author Declarations

This study was approved by the Medical Research and Ethics Committee (NMRR ID-25-02078-NSN), Ministry of Health Malaysia. The study utilized retrospective, de-identified patient data for evaluation of a clinical decision support tool. As the research involved analysis of existing clinical records without direct patient contact or intervention, informed consent was waived in accordance with MREC approval and national ethical guidelines.

## References

1. Kang J-S, Lee M (2009) Overview of Therapeutic Drug Monitoring. The Korean Journal of Internal Medicine 24:1. 10.3904/kjim.2009.24.1.1

2. Noman AA, Chakraborty A, Akter H, Binti RM (2023) Understanding Therapeutic drug monitoring (TDM) at a glance. International Journal of Medical Pharmacy and Drug Research 7:1. 10.22161/ijmpd.7.1.1

3. Kim B, Hwang S, Heo E, et al (2023) Evaluation of Vancomycin TDM Strategies: Prediction and Prevention of Kidney Injuries Based on Vancomycin TDM Results. Journal of Korean Medical Science. 10.3346/jkms.2023.38.e101

4. Pretorius E, Klinker H, Rosenkranz B (2011) The Role of Therapeutic Drug Monitoring in the Management of Patients With Human Immunodeficiency Virus Infection. Therapeutic Drug Monitoring 33:265. 10.1097/FTD.0b013e31821b42d1

5. Rybak MJ, Lê J, Lodise TP, et al (2020) Executive Summary: Therapeutic Monitoring of Vancomycin for Serious Methicillin-Resistant Staphylococcus aureus Infections: A Revised Consensus Guideline and Review of the American Society of Health-System Pharmacists, the Infectious Diseases Society of America, the Pediatric Infectious Diseases Society, and the Society of Infectious Diseases Pharmacists. Pharmacotherapy The Journal of Human Pharmacology and Drug Therapy 40:363. 10.1002/phar.2376

6. Gillett EM, Aleissa MM, Pearson JC, Solomon DA, Kubiak DW, Dionne B, Edrees HH, Okenla A, Chan BT (2024) Implementation of a Pharmacist-Driven Vancomycin Area Under the Concentration-Time Curve Monitoring Program Using Bayesian Modeling in Outpatient Parenteral Antimicrobial Therapy. Open Forum Infectious Diseases. 10.1093/ofid/ofae600

7. Alsowaida YS, Kubiak DW, Dionne B, Kovacevic MP, Pearson JC (2022) Vancomycin Area under the Concentration-Time Curve Estimation Using Bayesian Modeling versus First-Order Pharmacokinetic Equations: A Quasi-Experimental Study. Antibiotics 11:1239. 10.3390/antibiotics11091239

8. Abousheishaa AA, Sulaiman AH, Huri HZ, Kamis SF, Hamidi H, Ang WC, Zainal ZA, Shamsuddin N, Ng CG (2022) Psychiatric pharmaceutical care service across Malaysian hospitals: results from a cross-sectional study. BMC Health Services Research 22:321. 10.1186/s12913-022-07681-4

9. Park HY, Kim BY, Song JY, Seo KH, Lee SH, Choi S, Rhew K (2025) Effects of AUC-Based Vancomycin Therapeutic Drug Monitoring on AKI Incidence and Drug Utilization: A Propensity Score-Weighted Analysis. Journal of Clinical Medicine 14:1863. 10.3390/jcm14061863

10. Yager RC, Taylor N, Stocker SL, Day RO, Baysari M, Carland JE (2022) Would they accept it? An interview study to identify barriers and facilitators to user acceptance of a prescribing advice service. BMC Health Services Research 22:514. 10.1186/s12913-022-07927-1

11. Christof M, Armoundas AA (2025) Implications of integrating large language models into clinical decision making. Communications Medicine 5:490. 10.1038/s43856-025-01216-8

12. Bajwa J, Munir U, Nori A, Williams B (2021) Artificial intelligence in healthcare: transforming the practice of medicine. Future Healthcare Journal. 10.7861/fhj.2021-0095

13. Roosan D, Padua P, Khan R (2024) Effectiveness of ChatGPT in clinical pharmacy and the role of artificial intelligence in medication therapy management. J Am Pharm Assoc 64:422. 10.1016/j.japh.2023.11.023

14. Wang Z, Ong CLJ, Fu Z (2022) AI Models to Assist Vancomycin Dosage Titration. Frontiers in Pharmacology 13:801928. 10.3389/fphar.2022.801928

15. Liu X, Barreto EF, Dong Y, Liu C, Gao X, Tootooni MS, Song X, Kashani K (2023) Discrepancy between perceptions and acceptance of clinical decision support Systems: implementation of artificial intelligence for vancomycin dosing. BMC Medical Informatics and Decision Making. 10.1186/s12911-023-02254-9

16. Xu S, Most A, Chase A, Hedrick T, Murray B, Keats K, Smith S, Barreto E, Liu T, Sikora A (2024) Large language models management of complex medication regimens: a case-based evaluation. bioRxiv (Cold Spring Harbor Laboratory). 10.1101/2024.07.03.24309889

17. Ghasemiyeh P, Vazin A, Zand F, Haem E, Karimzadeh I, Azadi A, Masjedi M, Sabetian G, Nikandish R, Mohammadi-Samani S (2022) Pharmacokinetic assessment of vancomycin in critically ill patients and nephrotoxicity prediction using individualized pharmacokinetic parameters. Frontiers in Pharmacology. 10.3389/fphar.2022.912202

18. Onor IO, Neuliep A, Tran KA, Lambert J, Gillard C, Brakta F, Ezebuenyi M, James KSt, Okogbaa JI, Beyl RA (2020) Concordance of Vancomycin Population-Predicted Pharmacokinetics with Patient-Specific Pharmacokinetics in Adult Hospitalized Patients: A Case Series. Drugs in R&D 20:83. 10.1007/s40268-020-00298-0

19. Zhang N, Sun Z, Xie Y, Wu H, Li C (2024) The latest version ChatGPT powered by GPT-4o: what will it bring to the medical field? International Journal of Surgery 110:6018. 10.1097/JS9.0000000000001754

20. Islam R, Moushi OM (2024) GPT-4o: The Cutting-Edge Advancement in Multimodal LLM. 10.36227/techrxiv.171986596.65533294/v1

21. Clinical Pharmacy Working Committee (2019) Clinical Pharmacokinetics Pharmacy Handbook. Pharmacy Practice & Development Division, Ministry of Health Malaysia

22. Legg A, Devchand F, Gwee A, Sandaradura I, Lai T (2025) Safe and effective use of vancomycin. Australian Prescriber 48:54. 10.18773/austprescr.2025.013

23. American College of Clinical Pharmacy (2014) Standards of practice for clinical pharmacists. Pharmacotherapy 34:794. 10.1002/phar.1438

24. Shrout PE, Fleiss JL (1979) Intraclass correlations: Uses in assessing rater reliability. Psychological Bulletin 86:420. 10.1037/0033-2909.86.2.420

25. Koo TK, Li MY (2016) A Guideline of Selecting and Reporting Intraclass Correlation Coefficients for Reliability Research. Journal of Chiropractic Medicine 15:155. 10.1016/j.jcm.2016.02.012

26. Maus C, Rybacki S, Uhrmacher AM (2011) Rule-based multi-level modeling of cell biological systems. BMC Systems Biology 5:166. 10.1186/1752-0509-5-166

27. Papadopoulos P, Soflano M, Chaudy Y, Adejo W, Connolly T (2022) A systematic review of technologies and standards used in the development of rule-based clinical decision support systems. Health and Technology 12:713. 10.1007/s12553-022-00672-9

28. Wu A, Kuang K, Zhu M, Wang Y, Zheng Y, Han K, Li B, Chen G, Wu F, Zhang K (2024) Causality for Large Language Models. arXiv (Cornell University). 10.48550/arxiv.2410.15319

29. Landon SN, Savage T, Greysen SR, Bressman E (2025) Variation in Large Language Model Recommendations in Challenging Inpatient Management Scenarios. Journal of General Internal Medicine. 10.1007/s11606-025-09888-7

30. Dong C, Qiu X, Deng J, et al (2025) Comparative evaluation of large language models in delivering guideline-compliant recommendations for topical NSAID use in musculoskeletal pain: a multidimensional analysis. Clinical Rheumatology. 10.1007/s10067-025-07640-4

31. Reddy S (2024) Generative AI in healthcare: an implementation science informed translational path on application, integration and governance. Implementation Science. 10.1186/s13012-024-01357-9

32. Rybak MJ, Lê J, Lodise TP, et al (2020) Therapeutic monitoring of vancomycin for serious methicillin-resistant Staphylococcus aureus infections: A revised consensus guideline and review by the American Society of Health-System Pharmacists, the Infectious Diseases Society of America, the Pediatric Infectious Diseases Society, and the Society of Infectious Diseases Pharmacists. American Journal of Health-System Pharmacy 77:835. 10.1093/ajhp/zxaa036

33. Aydin S, Karabacak M, Vlachos V, Margetis K (2025) Navigating the potential and pitfalls of large language models in patient-centered medication guidance and self-decision support. Frontiers in Medicine 12:1527864. 10.3389/fmed.2025.1527864

34. Li G, Sun Y, Zhu L (2024) Application of machine learning combined with population pharmacokinetics to improve individual prediction of vancomycin clearance in simulated adult patients. Frontiers in Pharmacology. 10.3389/fphar.2024.1352113

35. Sessa M, Liang D, Khan AR, Külahçı M, Andersen M (2021) Artificial Intelligence in Pharmacoepidemiology: A Systematic Review. Part 2–Comparison of the Performance of Artificial Intelligence and Traditional Pharmacoepidemiological Techniques. Frontiers in Pharmacology. 10.3389/fphar.2020.568659

36. Rieke N, Hancox J, Li W, et al (2020) The future of digital health with federated learning. npj Digital Medicine. 10.1038/s41746-020-00323-1

