## Appendix B for "A Proof-of-Concept Study of a Clinical Decision Support System for Vancomycin Therapeutic Monitoring"

### Vancomycin TDM Assessment Rubric

#### Overview

This rubric evaluates the accuracy and clinical appropriateness of therapeutic drug monitoring (TDM) calculations and recommendations for vancomycin, comparing outputs from LLMs against pharmacist standards. Each domain is weighted to reflect its clinical importance.

#### Scoring System

- Score 0: Inadequate/Incorrect
- Score 1: Partially Correct
- Score 2: Fully Correct/Optimal

#### Domains and Criteria

##### A. Foundational Calculations (Weight: 2x)

**Accuracy Assessment**

| Item | Score 0 | Score 1 | Score 2 |
|---|---|---|---|
| Creatinine Clearance (CrCl) | Incorrect calculation or formula selection | Calculation within 10-20% of correct value or acceptable alternative equation | Calculation within <10% of correct value using appropriate equation |
| Ke (elimination rate const) | Incorrect calculation or formula | Calculation within 10-20% of correct value | Calculation within <10% of correct value |
| Half life (t1/2) | Incorrect calculation | Calculation within 10-20% of correct value | Calculation within <10% of correct value |
| Volume of distribution (Vd) | Incorrect calculation (>20% deviation) | Calculation within 10-20% of correct value | Calculation within <10% of correct value |
| CL (Clearance of drug) | Incorrect calculation (>20% deviation) | Calculation within 10-20% of correct value | Calculation within <10% of correct value |

**Clinical Appropriateness Assessment**

| Item | Score 0 | Score 1 | Score 2 |
|---|---|---|---|
| Equation selection for patient type | Inappropriate equation for patient population | Acceptable but not optimal equation for patient | Optimal equation selection for specific patient population |
| Patient-specific adjustments | No consideration of patient factors | Basic consideration of patient factors | Comprehensive consideration of all relevant patient factors |

#### B. Parameter Estimation & AUC (Weight: 3x)

**Accuracy Assessment**

| Item | Score 0 | Score 1 | Score 2 |
|---|---|---|---|
| Estimated Cmax | >20% deviation from expected value | 10-20% deviation from expected value | <10% deviation from expected value |
| Estimated Cmin | >20% deviation from expected value | 10-20% deviation from expected value | <10% deviation from expected value |
| Area under the curve (AUC) | >20% deviation or incorrect method | 10-20% deviation using acceptable method | <10% deviation using correct method |

**Clinical Appropriateness Assessment**

| Item | Score 0 | Score 1 | Score 2 |
|---|---|---|---|
| AUC/MIC ratio consideration | No consideration of AUC/MIC ratio | Basic consideration without specific targets | Appropriate target AUC/MIC ratio with clinical context |
| Method selection for estimation | Inappropriate method for clinical scenario | Acceptable but not optimal method | Optimal method selection based on available data |

#### C. Prospective Predictions (Weight: 3x)

##### Accuracy Assessment

| Item | Score 0 | Score 1 | Score 2 |
|---|---|---|---|
| New dose calculation | >20% deviation from established protocols | 10-20% deviation from established protocols | <10% deviation from established protocols |
| New Cmax prediction | >20% deviation from expected | 10-20% deviation from expected | <10% deviation from expected |
| New Cmin prediction | >20% deviation from expected | 10-20% deviation from expected | <10% deviation from expected |

##### Clinical Appropriateness Assessment

| Item | Score 0 | Score 1 | Score 2 |
|---|---|---|---|
| New dose recommendation | Potentially harmful or clinically inappropriate dose | Safe but suboptimal dose | Optimal dose considering all patient factors |
| Frequency adjustment | Inappropriate frequency for renal function | Acceptable frequency but not optimized | Optimal frequency based on patient parameters |
| Clinical context consideration | No consideration of clinical context | Basic consideration of clinical context | Comprehensive consideration of clinical context |

#### D. Timing Recommendations (Weight: 2x)

**Clinical Appropriateness Assessment**

| Item | Score 0 | Score 1 | Score 2 |
|---|---|---|---|
| Time to withhold (if needed) | No recommendation when needed or unsafe timeframe | Adequate timeframe but lacking patient-specific factors | Optimal timeframe with clear rationale and patient-specific factors |
| Time to repeat sample | Inappropriate timing or missing recommendation | Acceptable timing but suboptimal interval | Optimal timing with clear rationale based on PK principles |
| Urgency consideration | No consideration of clinical urgency | Basic consideration of urgency | Comprehensive consideration of urgency with clear guidance |

#### E. Interpretation & Core Recommendations (Weight: 5x)

**Accuracy Assessment**

| Item | Score 0 | Score 1 | Score 2 |
|---|---|---|---|
| Therapeutic range interpretation | Incorrect interpretation of levels | Basic interpretation with minor inaccuracies | Precise interpretation of levels relative to targets |
| Target identification | Incorrect target range | Acceptable but not optimal target range | Correct target range for specific indication |

**Clinical Appropriateness Assessment**

| Item | Score 0 | Score 1 | Score 2 |
|---|---|---|---|
| Dose suggestion | Incorrect or potentially harmful dosing | Safe but suboptimal dosing | Optimal dosing based on PK/PD principles and patient factors |
| Loading dose consideration | Failure to address loading dose when indicated | Basic loading dose recommendation | Optimal loading dose with clear rationale |
| Monitoring parameter | Missing or inappropriate monitoring plan | Basic monitoring plan | Comprehensive monitoring plan with clear timeframes |
| Special population considerations | No recognition of special population needs | Recognition without appropriate adjustments | Full recognition with appropriate adjustments |
| Clinical context integration | No integration of clinical context | Partial integration of clinical context | Full integration of all relevant clinical factors |

#### F. Documentation/Wording (Weight: 1x)

**Clinical Appropriateness Assessment**

| Item | Score 0 | Score 1 | Score 2 |
|---|---|---|---|
| Clarity of recommendation | Unclear, ambiguous, or confusing | Clear but lacking comprehensive details | Clear, concise, complete with all necessary details |
| Documentation completeness | Missing critical elements | Contains most necessary elements | Complete documentation with all necessary elements |
| Clinical reasoning explanation | No explanation of reasoning | Basic explanation of reasoning | Comprehensive explanation with clear rationale |

#### Overarching Safety Violations

| Violation Type | Assessment Category | Penalty |
|---|---|---|
| Dose exceeding maximum recommended daily dose | Clinical Appropriateness | -10 pts or automatic 0 case |
| Failure to recognize/address significant nephrotoxicity risk | Clinical Appropriateness | -10 pts or automatic 0 case |
| Incorrect frequency for renal function | Clinical Appropriateness | -10 pts or automatic 0 case |
| Failure to recommend therapeutic drug monitoring | Clinical Appropriateness | -10 pts or automatic 0 case |

#### Scoring Calculation

1. **Domain Score Calculation:**
    - Accuracy Assessment Score = Sum of accuracy item scores
    - Clinical Appropriateness Score = Sum of clinical appropriateness item scores
    - Total Domain Score = (Accuracy Score + Clinical Appropriateness Score) × Domain Weight
2. **Total Assessment Score:**
    - Raw Score = Sum of all domain scores
    - Final Score = Raw Score - Safety Violation Penalties
    - Percentage Score = (Final Score / Maximum Possible Score) × 100%
3. **Performance Classification:**
    - Excellent: 90-100%
    - Good: 80-89%
    - Acceptable: 70-79%
    - Needs Improvement: <70%

### Assessment Form

**Evaluator Name:** ___________________________
**Date of Evaluation:** ______________________
**Case/Scenario ID:** ________________________
**Output Source:** ☐ LLM ☐ Pharmacist

#### Domain Scores:

| Domain | Max Possible | Actual Score |
|---|---|---|
| A. Foundational Calculations (×2) | | |
| B. Parameter Estimation & AUC (×3) | | |
| C. Prospective Predictions (×3) | | |
| D. Timing Recommendations (×2) | | |
| E. Interpretation & Core Recommendations (×5) | | |
| F. Documentation/Wording (×1) | | |
| **Raw Score** | | |
| Safety Violation Penalties | | |
| **Final Score** | | |
| **Percentage Score** | | |

#### Comments:

#### Recommendations for Improvement:
