## Supplementary material for "A Proof-of-Concept Study of a Clinical Decision Support System for Vancomycin Therapeutic Monitoring": Apendix A: vancomycin_tdm_app.py

```
# -*- coding: utf-8 -*-
# TDM-AID (Vancomycin) with RAG + LLM Reasoning
# - Weight-choice rules for Cockcroft-Gault (TBW/IBW/AdjBW)
# - ADDED: Manual override for CrCl weight basis (Actual, Ideal, Adjusted)
# - Infusion-time guidance (>=60 min and <=10 mg/min)
# - Embeddings upgraded to text-embedding-3-small with robust fallback
#
# NOTE: This app preserves your calculation methods (pop-PK trough-only & Sawchuk-Zaske P+T).
#       Validate locally against MOH Clinical PK Handbook examples before production.

import os
import re
import json
import math
import faiss
import fitz  # PyMuPDF
import numpy as np
import pandas as pd
import streamlit as st
import tempfile
from pathlib import Path
from datetime import datetime, timedelta, time, date
import openai

# --- 1. OPENAI API KEY ---
openai.api_key = st.secrets.get("OPENAI_API_KEY", os.getenv("OPENAI_API_KEY"))
if not openai.api_key:
    st.error("OpenAI API key not found. Please set it in Streamlit secrets or as an environment variable.")

# --- 2. PAGE CONFIG ---
st.set_page_config(
    page_title="TDM-AID (Vancomycin) with RAG",
    page_icon="💊",
    layout="wide",
    initial_sidebar_state="expanded"
)

# --- 3. CSS ---
st.markdown("""
<style>
    .stContainer { padding: 10px 5px; }
    .stMetric > label { font-weight: bold; color: #555; }
    .stButton>button { width: 100%; }
    [data-testid="stVerticalBlock"] > [style*="flex-direction: column;"] > [data-testid="stVerticalBlock"] {
        border: 1px solid #e6e6e6; border-radius: 0.5rem; padding: 1rem; margin-bottom: 1rem; box-shadow: 0 1px 3px rgba(0,0,0,0.02);
    }
    h1 { color: #4F6BF2; padding-top: 10px; }
    .subtitle { color: #666; font-size: 0.9em; margin-top: -10px; margin-bottom: 20px; }
    .recommendation-dose { font-size: 1.5rem; font-weight: bold; color: #2A3C5D; margin-bottom: 5px; }
    .recommendation-description { font-size: 0.9rem; color: #666; }
    [data-testid="stHorizontalBlock"] > [data-testid="stVerticalBlock"]:first-child { display: flex; align-items: center; }
    .citation { background-color: #f0f7ff; border-left: 3px solid #4F6BF2; padding: 10px; margin: 10px 0; font-size: 0.9em; }
    .reasoning { background-color: #f9f9f9; border: 1px solid #ddd; border-radius: 5px; padding: 15px; margin-top: 15px; }
    .reasoning-title { font-weight: bold; margin-bottom: 10px; color: #2A3C5D; }
</style>
""", unsafe_allow_html=True)

# --- 4. RAG SYSTEM SETUP ---

class DocumentProcessor:
    def __init__(self, pdf_path):
        self.pdf_path = pdf_path
        self.chunks = []
        self.chunk_size = 1000
        self.overlap = 200

    def extract_text_from_pdf(self):
        try:
            doc = fitz.open(self.pdf_path)
            text = ""
            for page in doc:
                text += page.get_text()
            return text
        except Exception as e:
            st.error(f"Error extracting text from PDF: {e}")
            return None

    def chunk_text(self, text):
        if not text:
            return []
        # FIXED regex: use \s and \n (not escaped) inside raw string
        section_pattern = r'\n(?=[A-Z][A-Z\s]+(?:\n|:))'
        sections = re.split(section_pattern, text)

        chunks = []
        for section in sections:
            if not section.strip():
                continue
            if len(section) <= self.chunk_size:
                chunks.append(section.strip())
            else:
                words = section.split()
                current_chunk = []
                current_size = 0
                for word in words:
                    current_chunk.append(word)
                    current_size += len(word) + 1
                    if current_size >= self.chunk_size:
                        chunks.append(" ".join(current_chunk).strip())
                        overlap_size = 0
                        overlap_chunk = []
                        for w in reversed(current_chunk):
                            if overlap_size + len(w) + 1 <= self.overlap:
                                overlap_chunk.insert(0, w)
                                overlap_size += len(w) + 1
                            else:
                                break
                        current_chunk = overlap_chunk
                        current_size = overlap_size
                if current_chunk:
                    chunks.append(" ".join(current_chunk).strip())
        self.chunks = chunks
        return chunks

class RAGSystem:
    def __init__(self):
        self.embeddings = None
        self.chunks = None
        self.index = None
        self.is_initialized = False
        self.temp_dir = None
        self.embedding_model = None  # track active embedding model

    # ----- internal helpers -----
    def _pick_embedding_model(self):
        candidates = ["text-embedding-3-small", "text-embedding-3-large", "text-embedding-ada-002"]
        last_err = None
        for m in candidates:
            try:
                _ = openai.embeddings.create(model=m, input=["ok"])
                self.embedding_model = m
                return m
            except Exception as e:
                last_err = e
                continue
        raise RuntimeError(f"Unable to initialize any embedding model. Last error: {last_err}")

    def _ensure_model(self):
        if not self.embedding_model:
            self._pick_embedding_model()
        return self.embedding_model

    # ----- public API -----
    def embed_chunks(self, chunks):
        if not chunks:
            return None
        try:
            model = self._ensure_model()
            embeddings = []
            for i in range(0, len(chunks), 20):
                batch = chunks[i:i+20]
                resp = openai.embeddings.create(model=model, input=batch)
                batch_embeddings = [item.embedding for item in resp.data]
                embeddings.extend(batch_embeddings)
            self.embeddings = np.array(embeddings, dtype=np.float32)
            return self.embeddings
        except Exception as e:
            st.error(f"Error generating embeddings: {e}")
            return None

    def build_index(self):
        if self.embeddings is None:
            return False
        try:
            dim = self.embeddings.shape[1]
            self.index = faiss.IndexFlatL2(dim)
            self.index.add(self.embeddings)
            return True
        except Exception as e:
            st.error(f"Error building search index: {e}")
            return False

    def search(self, query, k=3):
        if not self.is_initialized or self.index is None or self.chunks is None:
            return []
        try:
            model = self._ensure_model()
            q = openai.embeddings.create(model=model, input=[query])
            q_embed = np.array([q.data[0].embedding], dtype=np.float32)
            distances, indices = self.index.search(q_embed, k)
            results = []
            for i, idx in enumerate(indices[0]):
                if 0 <= idx < len(self.chunks):
                    results.append({"text": self.chunks[idx], "score": float(distances[0][i])})
            return results
        except Exception as e:
            st.error(f"Error during search: {e}")
            return []

    def initialize_from_pdf(self, pdf_path):
        try:
            if not self.temp_dir:
                self.temp_dir = tempfile.TemporaryDirectory()
            processor = DocumentProcessor(pdf_path)
            text = processor.extract_text_from_pdf()
            if text:
                self.chunks = processor.chunk_text(text)
                self._ensure_model()
                self.embeddings = self.embed_chunks(self.chunks)
                success = self.build_index()
                self.is_initialized = success
                return success
            return False
        except Exception as e:
            st.error(f"Error initializing RAG system: {e}")
            return False

    def initialize_from_saved(self, chunks_path, embeddings_path, meta_path=None):
        try:
            with open(chunks_path, 'r') as f:
                self.chunks = json.load(f)
            self.embeddings = np.load(embeddings_path)
            if meta_path and Path(meta_path).exists():
                try:
                    with open(meta_path, 'r') as f:
                        meta = json.load(f)
                        self.embedding_model = meta.get("embedding_model")
                except Exception:
                    pass
            if not self.embedding_model:
                # Default to 3-small (1536 dims, same as ada-002)
                self.embedding_model = "text-embedding-3-small"
            success = self.build_index()
            self.is_initialized = success
            return success
        except Exception as e:
            st.error(f"Error loading saved RAG data: {e}")
            return False

    def save_data(self, chunks_path, embeddings_path, meta_path=None):
        try:
            with open(chunks_path, 'w') as f:
                json.dump(self.chunks, f)
            np.save(embeddings_path, self.embeddings)
            if meta_path:
                with open(meta_path, 'w') as f:
                    json.dump({"embedding_model": self.embedding_model or "text-embedding-3-small"}, f)
            return True
        except Exception as e:
            st.error(f"Error saving RAG data: {e}")
            return False

    def cleanup(self):
        if self.temp_dir:
            self.temp_dir.cleanup()

class LLMReasoner:
    def __init__(self):
        self.model = "gpt-4o"  # keep your original setting

    def generate_reasoning(self, patient_data, calculation_results, relevant_guidelines):
        try:
            guideline_context = "\n\n".join([item["text"] for item in relevant_guidelines]) if relevant_guidelines else ""
            patient_info = f"""
Patient Information:
- Age: {patient_data.get('age', 'N/A')} years
- Weight: {patient_data.get('weight', 'N/A')} kg
- Sex: {patient_data.get('sex', 'N/A')}
- SCr: {patient_data.get('scr', 'N/A')} µmol/L
- CrCl: {patient_data.get('crcl', 'N/A')} mL/min
- Clinical notes: {patient_data.get('clinical_notes', 'N/A')}
"""
            calc_info = ""
            if calculation_results:
                calc_info = "Calculation Results:\n"
                for key, value in calculation_results.items():
                    calc_info += f"- {key}: {value}\n"

            prompt = f"""You are a clinical pharmacist specialist in antimicrobial stewardship and pharmacokinetics. 
You are analyzing a vancomycin dosing regimen for a patient. Please provide your expert reasoning on the proposed dosing regimen.

{patient_info}

{calc_info}

Based on the relevant clinical guidelines, provide your reasoning:

{guideline_context}

Please provide:
1. An assessment of the current dosing and levels
2. Your reasoning on whether the calculated dose is appropriate
3. Any factors that might necessitate dose adjustment beyond the basic calculations
4. Specific rationale tied to the guidelines
5. Any monitoring recommendations

Format your response as a concise clinical consultation note. Include specific citations to the guidelines when making recommendations.
"""

            response = openai.chat.completions.create(
                model=self.model,
                messages=[
                    {"role": "system", "content": "You are a clinical pharmacist specialist providing expert consultation on vancomycin dosing."},
                    {"role": "user", "content": prompt}
                ],
                temperature=0.2,
                max_tokens=1500
            )
            return response.choices[0].message.content

        except Exception as e:
            st.error(f"Error generating LLM reasoning: {e}")
            return f"Unable to generate reasoning due to an error: {str(e)}"

# --- 5. RAG SINGLETON ---
@st.cache_resource
def get_rag_system():
    rag = RAGSystem()
    base_path = Path(".")
    chunks_path = base_path / "vanco_chunks.json"
    embeddings_path = base_path / "vanco_embeddings.npy"
    meta_path = base_path / "vanco_rag_meta.json"

    if chunks_path.exists() and embeddings_path.exists():
        if rag.initialize_from_saved(chunks_path, embeddings_path, meta_path):
            return rag

    pdf_path = base_path / "Vanco.pdf"
    if pdf_path.exists():
        if rag.initialize_from_pdf(pdf_path):
            rag.save_data(chunks_path, embeddings_path, meta_path)
            return rag

    st.error("Vancomycin guidelines PDF not found. Please make sure 'Vanco.pdf' is in the app directory.")
    return rag

# --- 6. HELPER FUNCTIONS ---

# Time helpers
def hours_diff(start: time, end: time) -> float:
    today = date.today()
    dt_start = datetime.combine(today, start)
    dt_end = datetime.combine(today, end)
    if dt_end <= dt_start:
        dt_end += timedelta(days=1)
    return (dt_end - dt_start).total_seconds() / 3600.0

def add_minutes_to_time(t: time, minutes: int) -> time:
    if not t:
        return None
    dt = datetime.combine(date.today(), t) + timedelta(minutes=minutes)
    return dt.time()

def format_hours_minutes(decimal_hours: float) -> str:
    if decimal_hours is None or decimal_hours < 0:
        return "Invalid time"
    total_minutes = int(round(decimal_hours * 60))
    h = total_minutes // 60
    m = total_minutes % 60
    if h > 0 and m > 0:
        return f"{h} hours {m} minutes"
    elif h > 0:
        return f"{h} hours"
    else:
        return f"{m} minutes"

# Weight-choice rules for CG CrCl
def ibw_devine(height_cm: float, is_female: bool) -> float:
    if not height_cm or height_cm <= 0:
        return None
    height_in = height_cm / 2.54
    base = 45.5 if is_female else 50.0
    return max(0.0, base + 2.3 * (height_in - 60))

def adjbw_func(ibw_kg: float, tbw_kg: float) -> float:
    return ibw_kg + 0.4 * (tbw_kg - ibw_kg)

def choose_weight_for_cg(tbw_kg: float, ibw_kg: float):
    if ibw_kg is None or tbw_kg is None or tbw_kg <= 0:
        return tbw_kg, "TBW (fallback)"
    ratio = tbw_kg / ibw_kg if ibw_kg > 0 else 1.0
    if tbw_kg < ibw_kg:
        return tbw_kg, "TBW (<IBW)"
    elif ratio >= 1.2:
        return adjbw_func(ibw_kg, tbw_kg), "AdjBW (>=120% IBW)"
    else:
        return ibw_kg, "IBW (100-120%)"

def calculate_crcl_cg(age, tbw_kg, scr_umol, is_female, height_cm, weight_choice="Auto-select"):
    if not all([age, tbw_kg, scr_umol]) or age <= 0 or tbw_kg <= 0 or scr_umol <= 0:
        return {'crcl': None, 'weight_used': None, 'weight_method': 'N/A', 'ibw': None, 'adjbw': None}

    # Always calculate IBW and AdjBW for reference and potential use
    ibw_kg = ibw_devine(height_cm, is_female) if height_cm else None
    adjbw_kg = adjbw_func(ibw_kg, tbw_kg) if ibw_kg is not None else None

    # Determine which weight to use based on user choice
    wt_used = None
    wt_method = ""

    if weight_choice == "Actual Body Weight":
        wt_used = tbw_kg
        wt_method = "TBW (manual)"
    elif weight_choice == "Ideal Body Weight":
        if ibw_kg is not None:
            wt_used = ibw_kg
            wt_method = "IBW (manual)"
        else:  # Fallback if height not provided
            wt_used, wt_method = choose_weight_for_cg(tbw_kg, ibw_kg)
            wt_method += " (IBW fallback)"
    elif weight_choice == "Adjusted Body Weight":
        if adjbw_kg is not None:
            wt_used = adjbw_kg
            wt_method = "AdjBW (manual)"
        else:  # Fallback if height not provided
            wt_used, wt_method = choose_weight_for_cg(tbw_kg, ibw_kg)
            wt_method += " (AdjBW fallback)"
    else:  # Default to "Auto-select"
        wt_used, wt_method = choose_weight_for_cg(tbw_kg, ibw_kg if ibw_kg else tbw_kg)

    # If for any reason weight is still not set, fallback to TBW
    if wt_used is None:
        wt_used = tbw_kg
        wt_method = "TBW (ultimate fallback)"

    scr_mgdl = scr_umol / 88.4
    if scr_mgdl <= 0:
        return {'crcl': None, 'weight_used': wt_used, 'weight_method': wt_method, 'ibw': ibw_kg, 'adjbw': adjbw_kg}

    crcl = ((140 - age) * wt_used) / (72 * scr_mgdl)
    if is_female:
        crcl *= 0.85
    crcl = max(0.0, crcl)
    return {
        'crcl': crcl,
        'weight_used': wt_used,
        'weight_method': wt_method,
        'ibw': ibw_kg,
        'adjbw': adjbw_kg
    }

# Target status + UI helpers
def check_target_status(value, target_range):
    if value is None or target_range is None:
        return "N/A"
    lower, upper = target_range
    try:
        value = float(value)
        if lower is not None and value < lower:
            return "BELOW TARGET"
        elif upper is not None and value > upper:
            return "ABOVE TARGET"
        elif lower is not None and upper is None and value < lower:
            return "BELOW TARGET"
        elif lower is not None:
            return "WITHIN TARGET"
        else:
            return "N/A"
    except (ValueError, TypeError):
        return "N/A"

def display_status(status_text):
    if status_text == "WITHIN TARGET":
        st.success(f"✓ {status_text}")
    elif status_text == "BELOW TARGET":
        st.warning(f"⚠️ {status_text}")
    elif status_text == "ABOVE TARGET":
        st.error(f"⚠️ {status_text}")
    else:
        st.info("Status N/A")

def display_level_indicator(label, value, target_range, unit):
    if value is None or target_range is None:
        st.metric(label=f"{label} ({unit})", value="N/A")
        return
    lower, upper = target_range
    status = check_target_status(value, target_range)
    value_str = f"{value:.1f}"
    delta_str, delta_color = None, "off"
    if status == "WITHIN TARGET":
        delta_str = "Target Met"
    elif status == "BELOW TARGET":
        delta_str, delta_color = f"↓ {lower}", "inverse"
    elif status == "ABOVE TARGET" and upper is not None:
        delta_str, delta_color = f"↑ {upper}", "inverse"
    elif status == "ABOVE TARGET" and upper is None:
        delta_str, delta_color = f"↑ {lower}", "normal"
    st.metric(label=f"{label} ({unit})", value=value_str, delta=delta_str, delta_color=delta_color)

# RAG query utility
def formulate_rag_query(patient_data, calculation_results, target_desc):
    query_parts = []
    if patient_data.get('crcl'):
        if patient_data['crcl'] < 30:
            query_parts.append("vancomycin dosing severe renal impairment")
        elif patient_data['crcl'] < 60:
            query_parts.append("vancomycin dosing moderate renal impairment")
    if calculation_results:
        # Simplified query since AUC is no longer primary
        if 'Trough Status' in calculation_results:
            if calculation_results['Trough Status'] == "BELOW TARGET":
                query_parts.append("vancomycin trough below target recommendations")
            elif calculation_results['Trough Status'] == "ABOVE TARGET":
                query_parts.append("vancomycin trough above target toxicity")
            else:
                query_parts.append("vancomycin trough within target monitoring")
    if "Empirical" in target_desc:
        query_parts.append("vancomycin empirical dosing guidelines trough 10-15")
    elif "Definitive/Severe" in target_desc:
        query_parts.append("vancomycin severe infection dosing guidelines trough 15-20")
    notes = (patient_data.get('clinical_notes') or "").lower()
    if "meningitis" in notes:
        query_parts.append("vancomycin CNS infection meningitis")
    if "endocarditis" in notes:
        query_parts.append("vancomycin endocarditis dosing")
    if "pneumonia" in notes:
        query_parts.append("vancomycin pneumonia dosing")
    if "dialysis" in notes or "hd" in notes or "crrt" in notes:
        query_parts.append("vancomycin renal replacement therapy dialysis")
    if "icu" in notes or "sepsis" in notes or "septic" in notes:
        query_parts.append("vancomycin critical care sepsis")
    if "obesity" in notes or "obese" in notes:
        query_parts.append("vancomycin dosing obesity")
    unique_parts = list(set(query_parts))
    if not unique_parts:
        unique_parts = ["vancomycin dosing guidelines monitoring recommendations"]
    return " ".join(unique_parts)

def display_rag_results(results):
    if not results:
        st.info("No relevant guideline sections found.")
        return
    st.subheader("Relevant Guideline Sections")
    for i, result in enumerate(results):
        text_block = result.get("text") or ""
        with st.expander(f"Guideline Section {i+1}"):
            st.markdown(f'<div class="citation">{text_block}</div>', unsafe_allow_html=True)

def display_llm_reasoning(reasoning_text):
    if not reasoning_text:
        st.info("No expert reasoning available.")
        return
    st.subheader("Expert Pharmacokinetic Analysis")
    st.markdown(f'<div class="reasoning"><p class="reasoning-title">Clinical Pharmacist Assessment</p>{reasoning_text}</div>', unsafe_allow_html=True)

def render_interpretation_st_peak_trough(trough_status, cmin_extrap, peak_status, cmax_extrap, auc_status, auc24, thalf, interval_h, new_dose, target_desc, pk_method):
    if not all([
        trough_status, cmin_extrap is not None, thalf is not None, interval_h, new_dose is not None, target_desc, auc24 is not None
    ]):
        st.warning("Interpretation cannot be generated due to missing calculation results.")
        return
        
    rec_action = "adjust"
    if "BELOW" in trough_status or "BELOW" in auc_status:
        rec_action = "increase"
    elif "ABOVE" in trough_status or "ABOVE" in auc_status:
        rec_action = "decrease"
    else:
        rec_action = "maintain"

    target_trough_str = "10-15 mg/L" if "Empirical" in target_desc else "15-20 mg/L"
    target_peak_str = "30-40 mg/L" # General target
    target_auc_str = "400-600 mg·h/L" if "Empirical" in target_desc else ">600 mg·h/L"


    with st.expander("View Detailed Interpretation", expanded=True):
        st.subheader("Assessment")
        assessment_text = f"""
- **Method:** {pk_method}
- **Extrapolated Trough (Cmin):** {cmin_extrap:.1f} mg/L ({trough_status.lower()})
- **Extrapolated Peak (Cmax):** {cmax_extrap:.1f} mg/L ({peak_status.lower()})
- **Calculated AUC₂₄:** {auc24:.1f} mg·h/L ({auc_status.lower()})
- **Calculated Half-life:** {f'{thalf:.1f}' if thalf is not None else 'N/A'} h
- **Current Interval:** q{interval_h}h (Interval is likely {'appropriate' if thalf is not None and interval_h >= thalf * 1.2 else 'potentially too long/short relative to half-life'})
"""
        st.markdown(assessment_text)
        st.subheader("Recommendation")
        recommendation_text = f"""
- **Action:** Consider **{rec_action}ing** the dose.
- **Suggested Regimen:** **{new_dose} mg q{interval_h}h**
- **Goal:** To achieve target trough ({target_trough_str}), peak ({target_peak_str}), and AUC ({target_auc_str}).
"""
        st.markdown(recommendation_text)
        st.subheader("Rationale")
        rationale_text = f"""
The recommendation aims to align the patient's trough, peak, and AUC levels with the selected therapeutic targets ({target_desc}).
The adjustment is based on the calculated pharmacokinetic parameters derived from the {pk_method.lower()} analysis.
"""
        st.markdown(rationale_text)
        st.subheader("Follow-up")
        followup_text = """
- Draw the next trough level before the 3rd or 4th dose of the *new* regimen to confirm target attainment.
- Continue to monitor renal function (e.g., SCr), clinical signs of infection, and potential vancomycin toxicity.
- Adjust therapy based on clinical response and subsequent levels.
"""
        st.markdown(followup_text)


# Infusion guidance helpers (>=60 min and <=10 mg/min), with auto-suggest support
def infusion_guidance(dose_mg: float):
    """
    Returns (message, minutes_by_rate_cap, minutes_recommended).
    Policy: infuse >=60 min AND do not exceed 10 mg/min.
    => recommended minutes = max(60, ceil(dose_mg / 10))
    """
    if not dose_mg or dose_mg <= 0:
        return ("", 0, 60)
    minutes_by_rate_cap = math.ceil(dose_mg / 10.0)
    minutes_recommended = max(60, minutes_by_rate_cap)
    msg = (f"Infusion guidance: infuse over >=60 minutes and not faster than 10 mg/min. "
           f"For {int(dose_mg)} mg, 10 mg/min ~ {minutes_by_rate_cap} min; "
           f"recommend >= {minutes_recommended} min.")
    return msg, minutes_by_rate_cap, minutes_recommended

# --- 7. MAIN APP ---

def main():
    rag_system = get_rag_system()
    llm_reasoner = LLMReasoner()

    # Header
    col_logo, col_title = st.columns([1, 5])
    with col_logo:
        try:
            st.image("logo.png", width=150)
        except Exception:
            st.markdown('<div style="font-size: 40px; margin-top: 10px;">💊</div>', unsafe_allow_html=True)
    with col_title:
        st.title("TDM-AID by HTAR")
        st.markdown('<p class="subtitle">VANCOMYCIN MODULE WITH RAG + LLM REASONING</p>', unsafe_allow_html=True)

    if not rag_system.is_initialized:
        st.warning("RAG system not initialized. Some advanced features may not be available.")

    # Sidebar
    with st.sidebar:
        st.header("⚙️ Patient Information")
        with st.container():
            col_pid, col_ward = st.columns(2)
            with col_pid:
                pid = st.text_input("Patient ID", placeholder="MRN12345")
            with col_ward:
                ward = st.text_input("Ward/Unit", placeholder="ICU")

        with st.container():
            col_age, col_wt = st.columns(2)
            with col_age:
                age = st.number_input("Age (years)", min_value=1, max_value=120, value=65, step=1)
            with col_wt:
                wt = st.number_input("Weight (kg)", min_value=1.0, max_value=300.0, value=70.0, step=0.5, format="%.1f")

            col_ht, col_scr, col_fem = st.columns([1,1,1])
            with col_ht:
                height_cm = st.number_input("Height (cm)", min_value=80.0, max_value=230.0, value=170.0, step=0.5, format="%.1f")
            with col_scr:
                scr_umol = st.number_input("SCr (µmol/L)", min_value=10.0, max_value=2000.0, value=88.0, step=1.0, format="%.0f", help="Serum Creatinine")
            with col_fem:
                st.markdown('<div style="height: 29px;"></div>', unsafe_allow_html=True)
                fem = st.checkbox("Female", value=False)
        
        # ADDED: Weight selection for CrCl
        crcl_weight_choice = st.selectbox(
            "CrCl Weight Basis",
            options=["Auto-select", "Actual Body Weight", "Ideal Body Weight", "Adjusted Body Weight"],
            index=0,
            help="Choose the weight for Cockcroft-Gault. 'Auto-select' uses standard criteria (TBW if <IBW, IBW if 100-120% IBW, AdjBW if >120% IBW)."
        )

        # CrCl Calculation (UPDATED CALL)
        crcl_data = calculate_crcl_cg(age, wt, scr_umol, fem, height_cm, crcl_weight_choice)
        crcl = crcl_data['crcl']
        if crcl is not None:
            st.metric(label="Estimated CrCl (mL/min)", value=f"{crcl:.1f}")
            st.caption(
                f"CG using {crcl_data['weight_method']}: {crcl_data['weight_used']:.1f} kg"
                + (f" | IBW {crcl_data['ibw']:.1f} kg" if crcl_data['ibw'] is not None else "")
                + (f" | AdjBW {crcl_data['adjbw']:.1f} kg" if crcl_data['adjbw'] is not None else "")
            )
        else:
            st.warning("Cannot calculate CrCl. Check Age, Weight, Height, or SCr.")

        # Targets
        with st.container():
            st.subheader("Therapeutic Target")
            target_level_desc = st.selectbox(
                "Select target level:",
                options=[
                    "Empirical (Target AUC24 400-600 mg·h/L; Trough ~10-15 mg/L)",
                    "Definitive/Severe (Target AUC24 >600 mg·h/L; Trough ~15-20 mg/L)"
                ],
                index=0,
                label_visibility="collapsed",
                help="Select the desired therapeutic range based on indication and severity."
            )
            if "Empirical" in target_level_desc:
                target_trough_range = (10.0, 15.0)
                target_auc_range = (400.0, 600.0) 
            else:
                target_trough_range = (15.0, 20.0)
                target_auc_range = (600.0, None)
            
            # General peak target, not for calculation but for status check
            target_peak_range = (30.0, 40.0)

        # Notes
        with st.container():
            st.subheader("Clinical Context")
            clinical_notes = st.text_area(
                "Clinical Notes",
                placeholder="Enter relevant clinical context (e.g., infection type, organ function, source control status...)",
                height=100,
                label_visibility="collapsed"
            )

        # Advanced toggles
        st.divider()
        st.subheader("Advanced Features")
        use_rag = st.toggle("Use RAG Guidelines", value=True, help="Retrieve relevant guideline sections for this case")
        use_llm = st.toggle("Use LLM Reasoning", value=True, help="Generate expert reasoning using GPT-4o")

    # Tabs
    tab1, tab2, tab3 = st.tabs(["Initial Dose", "Trough-Only Analysis", "Peak & Trough Analysis"])

    # --- INITIAL DOSE ---
    with tab1:
        st.header("Initial Loading Dose Calculator")
        st.caption("Calculate an appropriate one-time loading dose based on patient weight.")
        col1, col2 = st.columns([3,1])
        with col1:
            st.info("""
**Calculation Method:**
- Uses weight-based dosing (typically 20-35 mg/kg, commonly 25 mg/kg).
- Aims to rapidly achieve therapeutic concentrations.
- Dose is rounded to the nearest 250 mg increment.
- Note: Renal function primarily guides the maintenance dose interval, not the loading dose.
""")
        with col2:
            st.markdown('<div style="height: 50px;"></div>', unsafe_allow_html=True)
            calc_load_button = st.button("Calculate Loading Dose", key="calc_load")

        if calc_load_button:
            if wt is None or wt <= 0:
                st.error("Please enter a valid patient weight in the sidebar.")
            else:
                loading_dose_raw = wt * 25
                loading_dose_rounded = round(loading_dose_raw / 250) * 250
                max_loading_dose = 3000
                loading_dose_final = min(loading_dose_rounded, max_loading_dose)

                st.success(f"Recommended Initial Loading Dose: {loading_dose_final} mg (one-time)")
                st.caption(f"Calculated based on 25 mg/kg for {wt} kg weight, rounded to nearest 250 mg.")
                if loading_dose_final >= max_loading_dose:
                    st.warning(f"Loading dose capped at {max_loading_dose} mg.")

                # Infusion guidance
                guidance_msg, _, minutes_rec = infusion_guidance(loading_dose_final)
                st.info(guidance_msg)

                # RAG + LLM
                patient_data = {
                    'age': age, 'weight': wt, 'sex': 'Female' if fem else 'Male',
                    'scr': scr_umol, 'crcl': crcl, 'clinical_notes': clinical_notes
                }
                calculation_results = {
                    'Calculation Method': 'Initial Loading Dose',
                    'Calculated Dose (raw)': f"{loading_dose_raw:.1f} mg",
                    'Rounded Dose': f"{loading_dose_rounded} mg",
                    'Final Recommended Loading Dose': f"{loading_dose_final} mg (one-time)"
                }
                if use_rag and rag_system.is_initialized:
                    crcl_str = f"{crcl:.1f}" if crcl is not None else "NA"
                    query = f"vancomycin loading dose guidelines {wt}kg patient CrCl {crcl_str} {clinical_notes}"
                    results = rag_system.search(query, k=3)
                    if results:
                        display_rag_results(results)
                    if use_llm:
                        reasoning_text = llm_reasoner.generate_reasoning(patient_data, calculation_results, results)
                        display_llm_reasoning(reasoning_text)

                # Precompute strings for report
                weight_used_str = f"{crcl_data['weight_used']:.1f} kg" if crcl_data['weight_used'] is not None else "N/A"
                ibw_str = f"{crcl_data['ibw']:.1f} kg" if crcl_data['ibw'] is not None else "N/A"
                adjbw_str = f"{crcl_data['adjbw']:.1f} kg" if crcl_data['adjbw'] is not None else "N/A"

                report_data = f"""Vancomycin Initial Loading Dose Report
Date: {datetime.now().strftime('%Y-%m-%d %H:%M')}

Patient Information:
- ID: {pid if pid else 'N/A'}
- Ward: {ward if ward else 'N/A'}
- Age: {age} years
- Weight: {wt} kg
- Height: {height_cm} cm
- Sex: {'Female' if fem else 'Male'}
- SCr: {scr_umol} µmol/L
- Estimated CrCl: {f'{crcl:.1f}' if crcl is not None else 'N/A'} mL/min
- CrCl weight basis: {crcl_data['weight_method']} ({weight_used_str})
- IBW: {ibw_str}, AdjBW: {adjbw_str}

Loading Dose Calculation:
- Basis: 25 mg/kg
- Calculated Dose (raw): {loading_dose_raw:.1f} mg
- Rounded Dose: {loading_dose_rounded} mg
- Final Recommended Loading Dose: {loading_dose_final} mg (one-time)

Infusion Guidance: Infuse over >=60 minutes; do not exceed 10 mg/min (~ {minutes_rec} minutes recommended for this dose)

Clinical Notes:
{clinical_notes if clinical_notes else 'N/A'}

Disclaimer: For educational purposes only. Verify with clinical guidelines.
"""
                st.download_button(
                    label="Download Loading Dose Report",
                    data=report_data,
                    file_name=f"vanco_loading_dose_{pid if pid else 'report'}_{datetime.now().strftime('%Y%m%d_%H%M')}.txt",
                    mime="text/plain"
                )

    # --- TROUGH-ONLY ---
    with tab2:
        st.header("Trough-Only Analysis")
        st.caption("Estimate PK parameters and suggest dose adjustments based on a single steady-state trough level.")

        with st.form(key="trough_only_form"):
            col1, col2 = st.columns(2)
            with col1:
                st.subheader("Current Regimen")
                dose_current_to = st.number_input("Current Dose (mg)", min_value=250, step=250, value=1000, key="to_dose")
                interval_current_to = st.selectbox(
                    "Dosing Interval (hours)",
                    options=[6, 8, 12, 18, 24, 36, 48],
                    index=2,
                    format_func=lambda x: f"q{x}h",
                    key="to_interval"
                )
            with col2:
                st.subheader("Trough Level & Timing")
                trough_measured_to = st.number_input("Measured Trough (mg/L)", min_value=0.1, max_value=100.0, value=12.5, step=0.1, format="%.1f", key="to_trough")
                dose_time_to = st.time_input("Last Dose Given At", value=time(8, 0), step=timedelta(minutes=15), key="to_dose_time")
                sample_time_to = st.time_input("Trough Level Drawn At", value=time(19, 30), step=timedelta(minutes=15), key="to_sample_time")

                # Suggested infusion end for current dose
                _, _, minutes_rec_to = infusion_guidance(dose_current_to)
                st.caption(f"Suggested infusion end (info): {add_minutes_to_time(dose_time_to, minutes_rec_to).strftime('%H:%M')} (>=60 min and <=10 mg/min).")

            submitted_to = st.form_submit_button("Run Trough Analysis")

        if submitted_to:
            if not all([dose_current_to, interval_current_to, trough_measured_to, dose_time_to, sample_time_to]):
                st.error("Please ensure all inputs are provided.")
            elif crcl is None:
                st.error("Cannot perform analysis: CrCl could not be calculated. Check patient details in the sidebar.")
            else:
                timing_valid_to = True
                # MODIFIED LOGIC TO HANDLE PRE-DOSE TROUGH INPUT
                dt_dose = datetime.combine(date.today(), dose_time_to)
                dt_sample = datetime.combine(date.today(), sample_time_to)

                # If sample time is earlier on the same day as dose time, user likely entered the *next* dose time instead of the *last*.
                if dt_sample < dt_dose:
                    time_until_next_dose = (dt_dose - dt_sample).total_seconds() / 3600.0
                    
                    # Heuristic: if the time until dose is short (e.g., < 4 hours), assume it's a pre-dose level.
                    if 0 < time_until_next_dose < 4:
                         time_since_last_dose_h = interval_current_to - time_until_next_dose
                         st.info(f"Note: Trough sample appears to be a pre-dose level. Time since last dose was calculated as **{format_hours_minutes(time_since_last_dose_h)}** based on the q{interval_current_to}h interval, assuming the dose time you provided was for the *next* scheduled dose.")
                    else:
                        # Fall back to original logic which assumes next day.
                        time_since_last_dose_h = hours_diff(dose_time_to, sample_time_to)
                else:
                    time_since_last_dose_h = hours_diff(dose_time_to, sample_time_to)
                
                if time_since_last_dose_h <= 0:
                    st.error("Timing Error: Trough sample time must be after the last dose time.")
                    timing_valid_to = False
                elif time_since_last_dose_h >= interval_current_to:
                    st.warning(f"Timing Warning: Trough drawn {format_hours_minutes(time_since_last_dose_h)} after the dose, which is longer than the interval (q{interval_current_to}h). Ensure this timing is correct and represents a true trough.")

                if timing_valid_to:
                    with st.spinner("Analyzing trough level..."):
                        try:
                            # Population estimates
                            vd_est = 0.7 * wt  # TBW
                            ke_est = 0.00083 * crcl + 0.0044
                            if ke_est <= 0:
                                ke_est = 0.001
                            cl_est = ke_est * vd_est
                            thalf_est = math.log(2) / ke_est if ke_est > 0 else float('inf')
                            
                            # Trough-only AUC calculation remains for reference
                            auc_interval_est = dose_current_to / cl_est if cl_est > 0 else float('inf')
                            auc24_est = auc_interval_est * (24 / interval_current_to) if interval_current_to > 0 else float('inf')

                            # Recommendation based on trough proportion
                            target_trough_mid = (target_trough_range[0] + target_trough_range[1]) / 2
                            if trough_measured_to > 0:
                                new_dose_raw = (dose_current_to * target_trough_mid) / trough_measured_to
                                new_dose_rounded = round(new_dose_raw / 250) * 250
                            else:
                                new_dose_rounded = dose_current_to

                            trough_status = check_target_status(trough_measured_to, target_trough_range)
                            
                            pk_results = {
                                'Calculation Method': 'Trough-Only (Population Estimate)',
                                'Est. CrCl (mL/min)': f"{crcl:.1f}",
                                'Est. Vd (L)': f"{vd_est:.1f}",
                                'Est. Ke (h^-1)': f"{ke_est:.4f}",
                                'Est. CL (L/h)': f"{cl_est:.2f}",
                                'Est. t1/2 (h)': f"{thalf_est:.1f}",
                                'Est. AUC24 (mg·h/L)': f"{auc24_est:.1f}",
                                'Measured Trough (mg/L)': f"{trough_measured_to:.1f}",
                                'Trough Status': trough_status,
                                'Suggested New Dose': f"{new_dose_rounded} mg q{interval_current_to}h"
                            }

                            st.subheader("Analysis Results")
                            st.info(f"Trough drawn {format_hours_minutes(time_since_last_dose_h)} after the dose (Interval: q{interval_current_to}h). Calculations use population estimates based on patient demographics.")

                            res_col1, res_col2 = st.columns(2)
                            with res_col1:
                                st.subheader("Target Status")
                                display_level_indicator("Measured Trough", trough_measured_to, target_trough_range, "mg/L")
                                st.metric("Estimated AUC24", f"{auc24_est:.1f} mg·h/L", help="For reference only")
                            with res_col2:
                                st.subheader("Recommendation")
                                st.markdown(f'<p class="recommendation-dose">{new_dose_rounded} mg q{interval_current_to}h</p>', unsafe_allow_html=True)
                                st.markdown(f'<p class="recommendation-description">Suggested dose to achieve target {target_level_desc.split("(")[1].split(")")[0].strip()}.</p>', unsafe_allow_html=True)

                                # New dose infusion guidance
                                new_guidance_msg, _, minutes_rec_new = infusion_guidance(new_dose_rounded)
                                st.markdown(f"*{new_guidance_msg}*")

                            st.subheader("Estimated PK Parameters")
                            pk_col1, pk_col2, pk_col3 = st.columns(3)
                            with pk_col1:
                                st.metric("Est. Vd (L)", f"{vd_est:.1f}")
                                st.metric("Est. CL (L/h)", f"{cl_est:.2f}")
                            with pk_col2:
                                st.metric("Est. Ke (h^-1)", f"{ke_est:.4f}")
                                st.metric("Est. t1/2 (h)", f"{thalf_est:.1f}")
                            with pk_col3:
                                st.metric("CrCl (mL/min)", f"{crcl:.1f}")

                            patient_data = {
                                'age': age, 'weight': wt, 'sex': 'Female' if fem else 'Male',
                                'scr': scr_umol, 'crcl': crcl, 'clinical_notes': clinical_notes
                            }

                            if use_rag and rag_system.is_initialized:
                                query = formulate_rag_query(patient_data, pk_results, target_level_desc)
                                results = rag_system.search(query, k=3)
                                if results:
                                    display_rag_results(results)
                                if use_llm:
                                    reasoning_text = llm_reasoner.generate_reasoning(patient_data, pk_results, results)
                                    display_llm_reasoning(reasoning_text)

                            # Precompute strings for report
                            weight_used_str = f"{crcl_data['weight_used']:.1f} kg" if crcl_data['weight_used'] is not None else "N/A"
                            ibw_str = f"{crcl_data['ibw']:.1f} kg" if crcl_data['ibw'] is not None else "N/A"
                            adjbw_str = f"{crcl_data['adjbw']:.1f} kg" if crcl_data['adjbw'] is not None else "N/A"

                            report_data = f"""Vancomycin TDM Report (Trough-Only)
Date: {datetime.now().strftime('%Y-%m-%d %H:%M')}

Patient Information:
- ID: {pid if pid else 'N/A'}
- Ward: {ward if ward else 'N/A'}
- Age: {age} years
- Weight: {wt} kg
- Height: {height_cm} cm
- Sex: {'Female' if fem else 'Male'}
- SCr: {scr_umol} µmol/L
- Estimated CrCl: {f'{crcl:.1f}' if crcl is not None else 'N/A'} mL/min
- CrCl weight basis: {crcl_data['weight_method']} ({weight_used_str})
- IBW: {ibw_str}, AdjBW: {adjbw_str}

Current Regimen & Level:
- Dose: {dose_current_to} mg q{interval_current_to}h
- Last Dose Time: {dose_time_to.strftime('%H:%M')}
- Trough Sample Time: {sample_time_to.strftime('%H:%M')} ({format_hours_minutes(time_since_last_dose_h)} after dose)
- Measured Trough: {trough_measured_to:.1f} mg/L

Target: {target_level_desc}
- Target Trough: {target_trough_range[0]}-{target_trough_range[1]} mg/L

Analysis Results (Population Estimates):
{pd.Series(pk_results).to_string()}

Infusion Guidance (suggested new dose): Infuse over >=60 minutes; do not exceed 10 mg/min (~ {minutes_rec_new} minutes recommended)

Clinical Notes:
{clinical_notes if clinical_notes else 'N/A'}

Disclaimer: Trough-only analysis uses population estimates and has limitations. Individual PK may vary. Clinical correlation required.
"""
                            st.download_button(
                                label="Download Trough Analysis Report",
                                data=report_data,
                                file_name=f"vanco_trough_analysis_{pid if pid else 'report'}_{datetime.now().strftime('%Y%m%d_%H%M')}.txt",
                                mime="text/plain"
                            )

                        except Exception as e:
                            st.error(f"An error occurred during calculation: {e}")
                            st.exception(e)

    # --- PEAK & TROUGH ---
    with tab3:
        st.header("Peak & Trough Analysis")
        st.caption("Calculate individual PK parameters using paired peak and trough levels based on provided guide.")

        with st.form(key="peak_trough_form"):
            st.subheader("Regimen & Levels")
            col1, col2 = st.columns(2)
            with col1:
                dose_levels_pt = st.number_input("Dose Administered (mg)", min_value=250, step=250, value=1000, key="pt_dose")
                interval_pt = st.selectbox(
                    "Dosing Interval (hours)",
                    options=[6, 8, 12, 18, 24, 36, 48],
                    index=2,
                    format_func=lambda x: f"q{x}h",
                    key="pt_interval"
                )
            with col2:
                peak_measured_pt = st.number_input("Measured Peak (mg/L, Cpost)", min_value=0.1, max_value=200.0, value=30.0, step=0.1, format="%.1f", key="pt_peak")
                trough_measured_pt = st.number_input("Measured Trough (mg/L, Cpre)", min_value=0.1, max_value=100.0, value=10.0, step=0.1, format="%.1f", key="pt_trough")

            st.subheader("Timing Information")
            
            trough_sample_time_pt = st.time_input(
                "Trough Sample Time (t1, Pre-dose)",
                value=time(7, 30),
                step=timedelta(minutes=15),
                key="pt_trough_time"
            )
            infusion_start_time_pt = st.time_input(
                "Infusion Start Time",
                value=time(8, 0),
                step=timedelta(minutes=15),
                key="pt_inf_start"
            )
            infusion_end_time_pt = st.time_input(
                "Infusion End Time",
                value=time(9, 0),
                step=timedelta(minutes=15),
                key="pt_inf_end"
            )
            peak_sample_time_pt = st.time_input(
                "Peak Sample Time (t2, Post-dose)",
                value=time(10, 0),
                step=timedelta(minutes=15),
                key="pt_peak_time"
            )

            submitted_pt = st.form_submit_button("Run Peak & Trough Analysis")

        if submitted_pt:
            if not all([dose_levels_pt, interval_pt, peak_measured_pt, trough_measured_pt, infusion_start_time_pt, infusion_end_time_pt, peak_sample_time_pt, trough_sample_time_pt]):
                st.error("Please ensure all inputs are provided.")
            elif crcl is None:
                st.error("Cannot perform analysis: CrCl could not be calculated. Check patient details in the sidebar.")
            elif peak_measured_pt <= trough_measured_pt:
                st.error("Input Error: Peak level must be higher than trough level.")
            else:
                # --- ALIGNED WITH GUIDE FORMULAS ---
                
                # Time variables from guide
                t1 = trough_sample_time_pt
                t2 = peak_sample_time_pt
                T = float(interval_pt) # Dosing interval
                
                # Time differences in hours
                infusion_duration_h = hours_diff(infusion_start_time_pt, infusion_end_time_pt) # This is t' from the AUC formula
                time_between_samples = hours_diff(t1, t2)
                t_prime = hours_diff(infusion_end_time_pt, t2) # t' from Cmax guide
                
                timing_valid_pt = True
                if time_between_samples <= 0:
                    st.error("Timing Error: Peak sample time (t2) must be after trough sample time (t1).")
                    timing_valid_pt = False
                if T <= time_between_samples:
                    st.error("Timing Error: Dosing interval (T) must be longer than the time between samples (t2-t1).")
                    timing_valid_pt = False
                if t_prime < 0:
                    st.error("Timing Error: Peak sample time (t2) must be after infusion end time.")
                    timing_valid_pt = False

                if timing_valid_pt:
                    with st.spinner("Analyzing peak and trough levels..."):
                        try:
                            # a) Ke Calculation
                            ke_denominator = T - time_between_samples
                            if ke_denominator <= 0:
                                raise ValueError("Ke denominator is zero or negative. Check Dosing Interval and sample times.")
                            # Use ln(peak) - ln(trough) to get a positive Ke
                            ke_ind = math.log(peak_measured_pt / trough_measured_pt) / ke_denominator
                            if ke_ind <= 0:
                                raise ValueError("Calculated Ke is non-positive. Check levels and times.")

                            # b) Half-life
                            thalf_ind = 0.693 / ke_ind

                            # c) Cmax Calculation
                            cmax_extrap = peak_measured_pt * math.exp(ke_ind * t_prime)
                            
                            # d) Cmin Calculation (MODIFIED)
                            # METHOD 1: Extrapolate from Cmax down (less reliable due to distribution phase effects)
                            cmin_from_peak = cmax_extrap * math.exp(-ke_ind * T)
                            
                            # METHOD 2: Extrapolate from measured trough (Cpre) forward (more reliable, now primary method)
                            time_trough_to_dose = hours_diff(trough_sample_time_pt, infusion_start_time_pt)
                            if time_trough_to_dose < 0:
                                st.warning("Timing Warning: Trough sample drawn after infusion started. Cmin (from Trough) may be inaccurate.")
                                time_trough_to_dose = 0
                            cmin_extrap = trough_measured_pt * math.exp(-ke_ind * time_trough_to_dose)


                            # e) Vd Calculation (L/kg)
                            vd_denominator = (wt * cmax_extrap * (1 - math.exp(-ke_ind * T)))
                            if vd_denominator == 0:
                                raise ValueError("Vd denominator is zero. Cannot calculate.")
                            vd_ind_per_kg = dose_levels_pt / vd_denominator
                            vd_ind_total = vd_ind_per_kg * wt # Total Vd in L
                            
                            # AUC Calculation using Trapezoidal Method
                            auc_inf = infusion_duration_h * (cmin_extrap + cmax_extrap) / 2
                            auc_elim = (cmax_extrap - cmin_extrap) / ke_ind if ke_ind > 0 else 0
                            auc_interval = auc_inf + auc_elim
                            auc24_trap = auc_interval * (24 / T) if T > 0 else 0

                            # f & g) New Dose Recommendation
                            # Define a target Cmax based on the selected trough range
                            target_cmax = 40.0 if "Definitive/Severe" in target_level_desc else 35.0

                            new_dose_raw = target_cmax * vd_ind_total * (1 - math.exp(-ke_ind * T))
                            new_dose_rounded = round(new_dose_raw / 250) * 250
                            
                            # Predict new Cmin with the new dose
                            if (vd_ind_total > 0) and (1-math.exp(-ke_ind * T) > 0):
                                expected_cmax_new_dose = new_dose_rounded / (vd_ind_total * (1 - math.exp(-ke_ind * T)))
                                expected_cmin_new_dose = expected_cmax_new_dose * math.exp(-ke_ind * T)
                            else:
                                expected_cmax_new_dose = 0
                                expected_cmin_new_dose = 0


                            trough_status = check_target_status(cmin_extrap, target_trough_range)
                            peak_status = check_target_status(cmax_extrap, target_peak_range)
                            auc_status = check_target_status(auc24_trap, target_auc_range)

                            pk_results = {
                                'Calculation Method': 'Peak & Trough (Guide Formula)',
                                'Individual Vd (L/kg)': f"{vd_ind_per_kg:.2f}",
                                'Individual Ke (h^-1)': f"{ke_ind:.4f}",
                                'Individual t1/2 (h)': f"{thalf_ind:.1f}",
                                'Extrapolated Cmax (mg/L)': f"{cmax_extrap:.1f}",
                                'Extrapolated Cmin (from Trough)': f"{cmin_extrap:.1f}",
                                'Extrapolated Cmin (from Peak, for reference)': f"{cmin_from_peak:.1f}",
                                'AUC24 (Trapezoidal)': f"{auc24_trap:.1f}",
                                'Trough Status': trough_status,
                                'Peak Status': peak_status,
                                'AUC Status': auc_status,
                                'Suggested New Dose': f"{new_dose_rounded} mg q{interval_pt}h",
                                'Predicted Cmin with New Dose': f"{expected_cmin_new_dose:.1f} mg/L"
                            }

                            st.subheader("Analysis Results")
                            analysis_summary_string = (
                                f"Dosing Interval (T): {T}h, "
                                f"Time Between Samples (t2-t1): {format_hours_minutes(time_between_samples)}, "
                                f"Time Infusion End -> Peak (t'): {format_hours_minutes(t_prime)}"
                            )
                            st.info(analysis_summary_string)
                            st.caption(f"Primary Cmin is calculated by extrapolating the measured trough forward, as this is generally more reliable.")


                            res_col1, res_col2 = st.columns(2)
                            with res_col1:
                                st.subheader("Target Status")
                                display_level_indicator("Extrapolated Trough (Cmin)", cmin_extrap, target_trough_range, "mg/L")
                                display_level_indicator("Extrapolated Peak (Cmax)", cmax_extrap, target_peak_range, "mg/L")
                                display_level_indicator("Calculated AUC24", auc24_trap, target_auc_range, "mg·h/L")
                            with res_col2:
                                st.subheader("Recommendation")
                                st.markdown(f'<p class="recommendation-dose">{new_dose_rounded} mg q{interval_pt}h</p>', unsafe_allow_html=True)
                                st.markdown(f'<p class="recommendation-description">Suggested dose to achieve target Cmax of ~{target_cmax} mg/L. Predicted Cmin: {expected_cmin_new_dose:.1f} mg/L.</p>', unsafe_allow_html=True)
                                new_guidance_msg, _, minutes_rec_new = infusion_guidance(new_dose_rounded)
                                st.markdown(f"*{new_guidance_msg}*")

                            st.subheader("Individual PK Parameters")
                            pk_col1, pk_col2, pk_col3 = st.columns(3)
                            with pk_col1:
                                st.metric("Individual Vd (L/kg)", f"{vd_ind_per_kg:.2f}")
                                st.metric("Individual Ke (h^-1)", f"{ke_ind:.4f}")
                            with pk_col2:
                                st.metric("Individual t1/2 (h)", f"{thalf_ind:.1f}")
                                st.metric("Extrap. Cmax (mg/L)", f"{cmax_extrap:.1f}")
                            with pk_col3:
                                st.metric("Extrap. Cmin (mg/L)", f"{cmin_extrap:.1f}")
                                st.metric("Total Vd (L)", f"{vd_ind_total:.1f}")

                            render_interpretation_st_peak_trough(
                                trough_status=trough_status,
                                cmin_extrap=cmin_extrap,
                                peak_status=peak_status,
                                cmax_extrap=cmax_extrap,
                                auc_status=auc_status,
                                auc24=auc24_trap,
                                thalf=thalf_ind,
                                interval_h=interval_pt,
                                new_dose=new_dose_rounded,
                                target_desc=target_level_desc,
                                pk_method="Peak & Trough (Guide Formula)"
                            )

                            patient_data = {
                                'age': age, 'weight': wt, 'sex': 'Female' if fem else 'Male',
                                'scr': scr_umol, 'crcl': crcl, 'clinical_notes': clinical_notes
                            }

                            if use_rag and rag_system.is_initialized:
                                query = formulate_rag_query(patient_data, pk_results, target_level_desc)
                                results = rag_system.search(query, k=3)
                                if results:
                                    display_rag_results(results)
                                if use_llm:
                                    reasoning_text = llm_reasoner.generate_reasoning(patient_data, pk_results, results)
                                    display_llm_reasoning(reasoning_text)

                            # Precompute strings for report
                            weight_used_str = f"{crcl_data['weight_used']:.1f} kg" if crcl_data['weight_used'] is not None else "N/A"
                            ibw_str = f"{crcl_data['ibw']:.1f} kg" if crcl_data['ibw'] is not None else "N/A"
                            adjbw_str = f"{crcl_data['adjbw']:.1f} kg" if crcl_data['adjbw'] is not None else "N/A"

                            report_data = f"""Vancomycin TDM Report (Peak & Trough)
Date: {datetime.now().strftime('%Y-%m-%d %H:%M')}

Patient Information:
- ID: {pid if pid else 'N/A'}
- Ward: {ward if ward else 'N/A'}
- Age: {age} years
- Weight: {wt} kg
- Height: {height_cm} cm
- Sex: {'Female' if fem else 'Male'}
- SCr: {scr_umol} µmol/L
- Estimated CrCl: {f'{crcl:.1f}' if crcl is not None else 'N/A'} mL/min
- CrCl weight basis: {crcl_data['weight_method']} ({weight_used_str})
- IBW: {ibw_str}, AdjBW: {adjbw_str}

Regimen, Levels & Timing:
- Dose: {dose_levels_pt} mg q{interval_pt}h
- Trough Sample Time (t1): {t1.strftime('%H:%M')}
- Peak Sample Time (t2): {t2.strftime('%H:%M')}
- Measured Peak: {peak_measured_pt:.1f} mg/L
- Measured Trough: {trough_measured_pt:.1f} mg/L

Target: {target_level_desc}

Analysis Results (Guide Formulas):
{pd.Series(pk_results).to_string()}

Infusion Guidance (suggested new dose): Infuse over >=60 minutes; do not exceed 10 mg/min (~ {minutes_rec_new} minutes recommended)

Clinical Notes:
{clinical_notes if clinical_notes else 'N/A'}

Disclaimer: For educational purposes only. Verify calculations and clinical correlation.
"""
                            st.download_button(
                                label="Download Peak & Trough Analysis Report",
                                data=report_data,
                                file_name=f"vanco_peak_trough_analysis_{pid if pid else 'report'}_{datetime.now().strftime('%Y%m%d_%H%M')}.txt",
                                mime="text/plain"
                            )

                        except ValueError as ve:
                            st.error(f"Calculation Error: {ve}. Please check input values and timings.")
                        except Exception as e:
                            st.error(f"An unexpected error occurred during calculation: {e}")
                            st.exception(e)

    # Footer
    st.markdown("---")
    st.caption("""
Disclaimer: This tool is intended for educational and informational purposes only and does not constitute medical advice.
Calculations are based on standard pharmacokinetic principles but may need adjustment based on individual patient factors and clinical context.
Always consult with qualified healthcare professionals and local guidelines for clinical decision-making.
Reference: Basic Clinical Pharmacokinetics (6th Ed.), Clinical Pharmacokinetics Pharmacy Handbook (2nd Ed.), ASHP/IDSA Vancomycin Guidelines.

Developed by Dr. Fahmi Hassan, Enhanced with RAG and LLM reasoning.
""")

    try:
        rag_system.cleanup()
    except Exception:
        pass

if __name__ == "__main__":
    main()
```
